# Nuclear Genome Amplification of Antibody VDJ Regions in Wastewater

**DOI:** 10.64898/2026.09.21.26363604

**Authors:** D. Chirman, J.C Clark, S. Bel Rhali, E Boerwinkle, A.W. Maresso

## Abstract

Wastewater-based epidemiology (WBE) has become a valuable tool to monitor pathogens from community and municipal sources. Whereas the detection of a pathogen in wastewater can help public health agencies understand what is present in a community, it cannot provide insight into a population’s preexisting immunity. Genomic rearrangement of the VDJ region of the human genome generates antibody diversity, a critical step in human immunity. We hypothesized that human antibody genes could be amplified from wastewater and sequenced, providing a snapshot of a population’s antibody repertoire. Here, we report the successful amplification and sequencing of human immunoglobulin heavy and light chain variable regions from wastewater. We observed a diverse range of sequence polymorphisms in these reads, suggesting the presence of distinct antibody sequences from various clonal lineages. The sequences aligned to conserved structural regions as well as the more variable complementarity-determining regions (CDRs). Additionally, we aligned the wastewater reads to two databases of antibodies with known functions. The wastewater reads encoded the conserved CDRs1 and 2, but diverged at the CDR3 sites, which determine epitope binding. Collectively, these findings serve as proof of principle that wastewater analysis may extend beyond pathogen detection to the characterization of population immunity and human genetics more broadly.

## Introduction

Wastewater-based epidemiology (WBE) has expanded rapidly in both research and application. Tracking SARS-CoV-2 in wastewater accurately predicts infections and hospital admissions^1–5^ and monitor the evolution of new variants^6,7^. Because samples are collected at community catchment sites, WBE resolves viral signal to specific neighborhoods within a metropolis^8^, and even buildings^9,10^. This success prompted state and local agencies to deploy WBE against recent monkeypox and polio outbreaks as case numbers rose^11–16^. Many entities now routinely monitor for pathogens, including those using sequencing to monitor the pathome (TexWEB Texas Wastewater and Environmental Biomonitoring)^17,18^, programs that report on seasonal viruses using sensitive techniques such as PCR (WastewaterSCAN/Verily)^19,20^, and commercial entities such as Biobot, which also monitors for drug substances. United States Centers for Disease Control and Prevention has also established the National Wastewater Surveillance System (NWSS) to track respiratory viruses^21^.

One key determinant of whether a pathogen will cause an epidemic is the existence of prior immunity. If a critical threshold of the population does have a reactive adaptive immune response against that pathogen, it cannot sufficiently spread between naïve hosts to cause an epidemic. We therefore hypothesized that it is possible to monitor the immunological status of a population based on its repertoire of antibodies encoded in DNA present in wastewater. A recently published study and pre-print have both shown that antibodies can be isolated from wastewater^22,23^, including antibodies specific for respiratory pathogens SARS-CoV-2, respiratory syncytial virus, and influenza A virus. Moreover, these antibodies increased in abundance during months of greater viral transmission and vaccine uptake^23^. However, there are no reports of a genomic analysis of antibody repertoires from wastewater samples even though sequencing the genetic determinants of antibody structure are possible, high-throughput and unbiased. Knowing the antibody rearrangement repertoires from a population may facilitate (1) understanding of the total diversity of antibody possibilities in a population, as opposed to individual, level; (2) determination of the frequency or abundance of important antibody-antigen pairs, especially if matched to a wastewater analysis of cognate pathogens in the same sample; (3) and a cataloguing of the ability of that population to produce neutralizing or functional antibodies against important and well-known seasonal pathogens, possibly to assess the degree of herd immunity against such pathogens. Finally, the sequencing of antibody genes may allow for the discovery of new antibodies against previously undiscovered antigens, which may inform on possible vaccine design or antibody synthesis for treatments (e.g. library generation to identify new antibody therapy candidates)^24^.

Here, we report the successful amplification and sequencing of the immunoglobulin heavy chain variable region (IGHV) and the immunoglobulin kappa and lambda variable clusters (IGKV and IGLV) from wastewater. These sequences undergo a process known as V(D)J recombination by combining with diversity and/or joining gene segments to produce a diverse repertoire of antibody heavy and light chains, respectively. With these sequences, we were able to characterize the allele utilization of the IGHV and IGKV genes in amplified V(D)J sequences for each wastewater sample and align them to a database of antibodies with known functions. This report provides proof-of-principle that antibody repertoires can be amplified, sequenced, and annotated from wastewater.

## Methods

### 1. Wastewater collection

Influent of unprocessed wastewater was obtained from three distinct wastewater catchment sites across separate cities and regions in the state of Texas. The identities of these sites are kept anonymous given the sensitive nature of the gene sequences being reported here. Instead, the sample collection sites are referred to as Site A, Site B, and Site C, respectively, based on the order in which they were processed. Samples A-C had collection dates on 8 January 2024, 27 November 2023, and 15 January 2024, respectively. For each sample, roughly 300 mL of influent sewage was collected from 24 hour composite samplers and bottled. Bottles were decontaminated by surface cleaning with 10% bleach prior to bagging and shipment to Baylor College of Medicine on ice. Obtained samples are stored at 4°C.

### 2. DNA extraction and purification

Wastewater samples were filtered through a 0.22 um vacuum filter for isolation of concentrated biosolids. The filter retentate was collected by resuspension in 25 mL PBS and re-pelleted by centrifugation at 1,000 x g for 20 minutes. Supernatant was discarded and the pellet was weighed to quantify the amount of biosolids obtained. Samples A-C yielded 0.8 g, 0.25 g, and 0.12 g, respectively. These pellets were resuspended in 1 mL PBS and lysed with lysis buffer from the Promega Magazorb DNA Extraction Kit (Promega). The remainder of the DNA extraction was carried out using the kit’s manufacturer instructions. Briefly, DNA was bound to the kit’s magnetic beads by mixing with binding buffer. Beads were then magnetically sedimented, washed, and the bound DNA was eluted. Eluent of purified DNA was cleaned up with the QIAQuick PCR and Gel Cleanup Kit (Qiagen) according to manufacturer instructions. DNA purity and concentration were assessed by Nanodrop (Thermo Scientific). Purified DNA extracts are stored at −20°C.

As a positive control, DNA extracts were also obtained from a sample of 50 uL of peripheral blood mononuclear cells (PBMCs; Charles River Laboratories) suspended in 150 uL PBS (5 x 10^5^ total cells). An additional positive control was included by spiking the same concentration of PBMCs (5 x 10^5^ total cells in 200 uL) into a separate sample of wastewater resuspension from Site A. This control was included to ensure that the PCR assay could productively amplify the target gene even in the presence of residual contaminants from the undefined, complex mixture of wastewater used for our samples. DNA was extracted from these samples using the same reagents and protocol as described above.

### 3. PCR amplification of gene targets

Three gene targets were amplified by nested multiplex polymerase chain reaction (PCR) at the following loci: the immunoglobulin heavy chain variable region (IGHV), the immunoglobulin kappa variable cluster (IGKV), and the immunoglobulin lambda variable cluster (IGLV). This was done using the Qiagen multiplex PCR kit according to the manufacturer’s standard protocol. Template DNA was obtained from the purified DNA extraction from each wastewater catchment site. Template DNA for the positive controls included the PBMC DNA extract and the DNA extract of wastewater spiked with PBMCs, described above. A negative control with no template DNA (eluent only) was included. Primers (specified in **Supplemental Table 1**) were selected on the basis of a previously published protocol for VDJ amplification and sequencing^25^. Primers were used at a final concentration of 0.1 uM to accommodate the large number of primer targets. The PCR thermocycling protocol used for both nested reactions is shown in **Supplemental Table 2**. These settings were consistent with the recommended protocol, but the annealing time was increased to 3 minutes because of the suspected low abundance of target DNA and the large number of amplicon targets.

**Supplemental Table 1:**
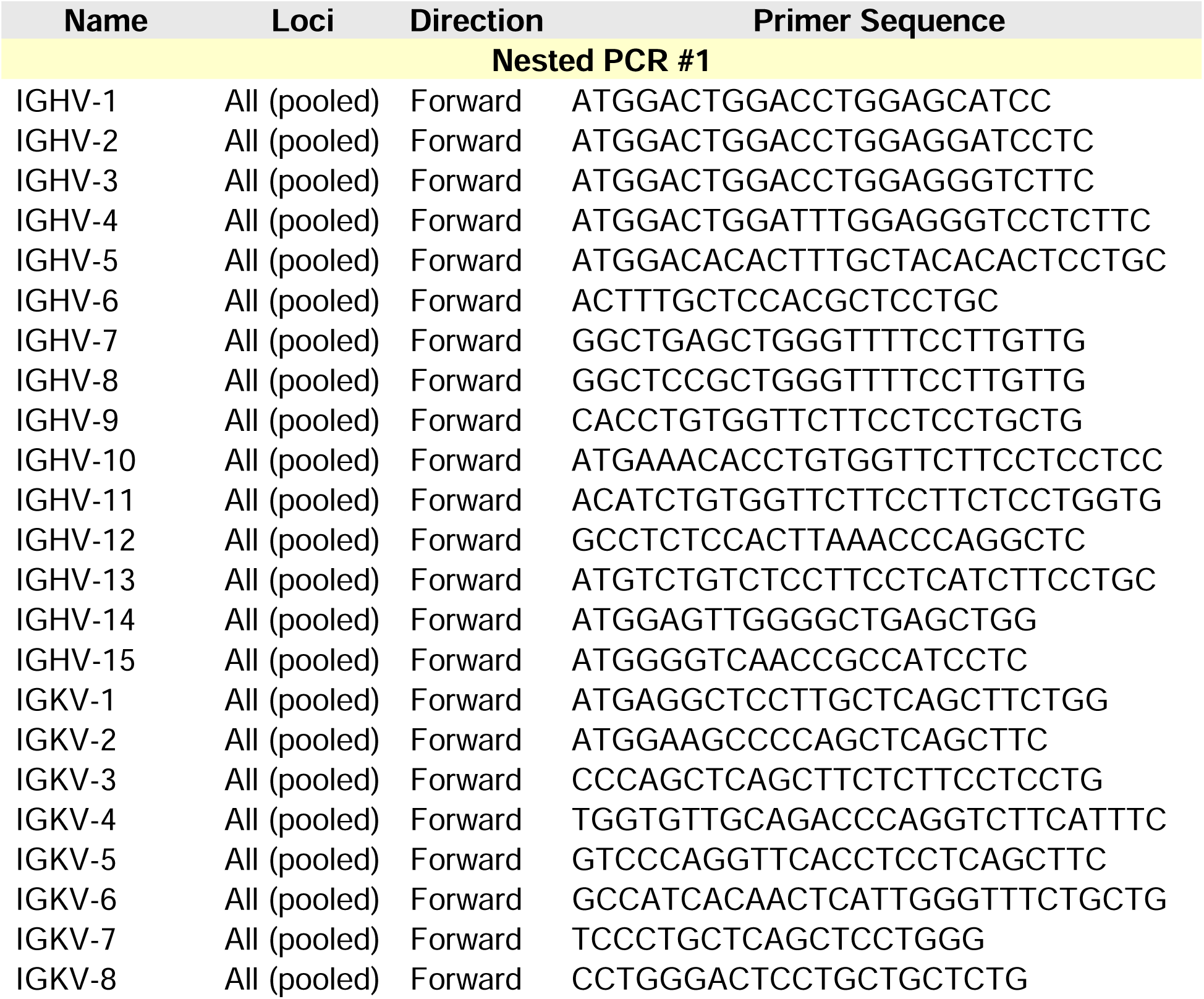
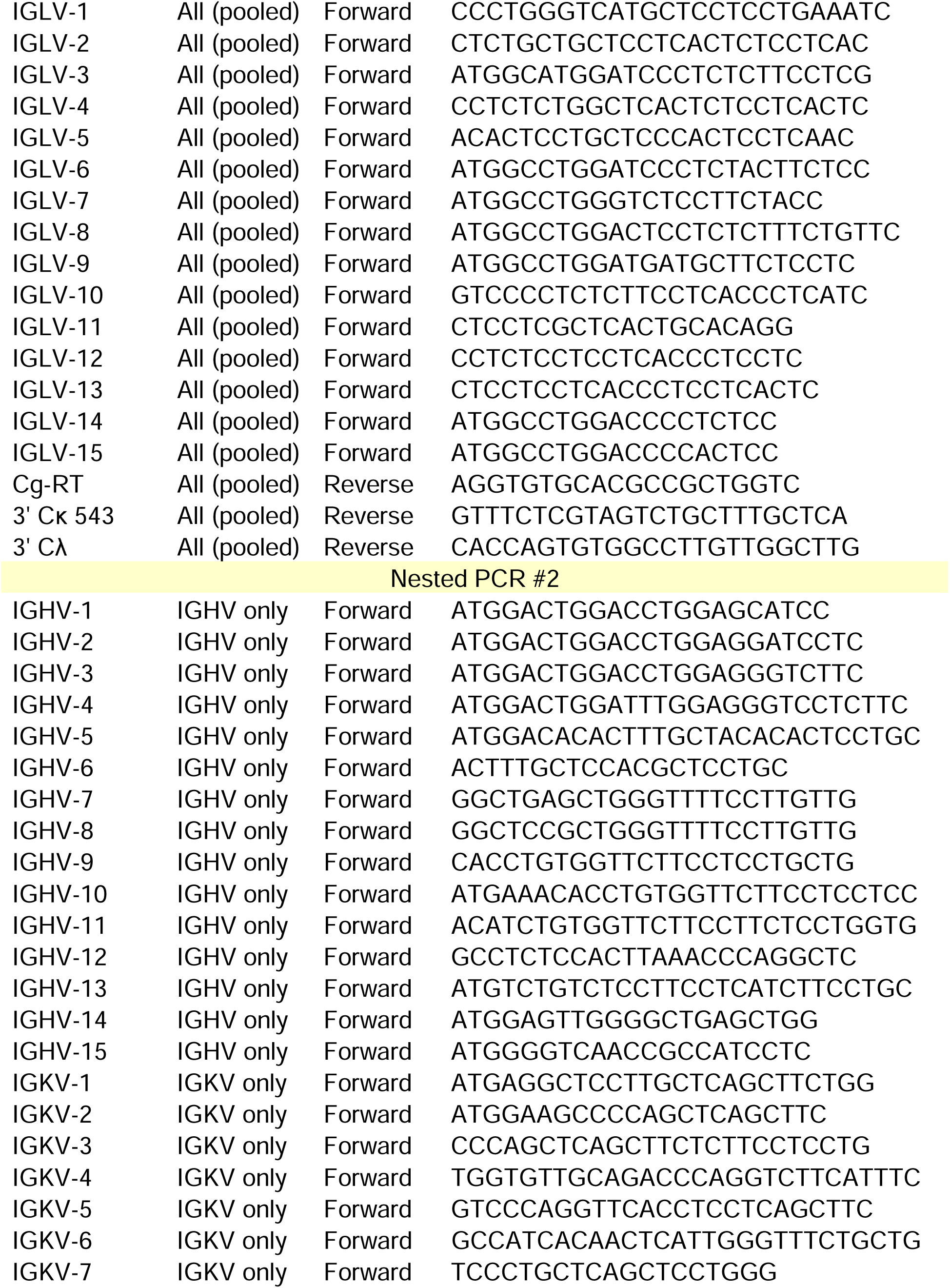
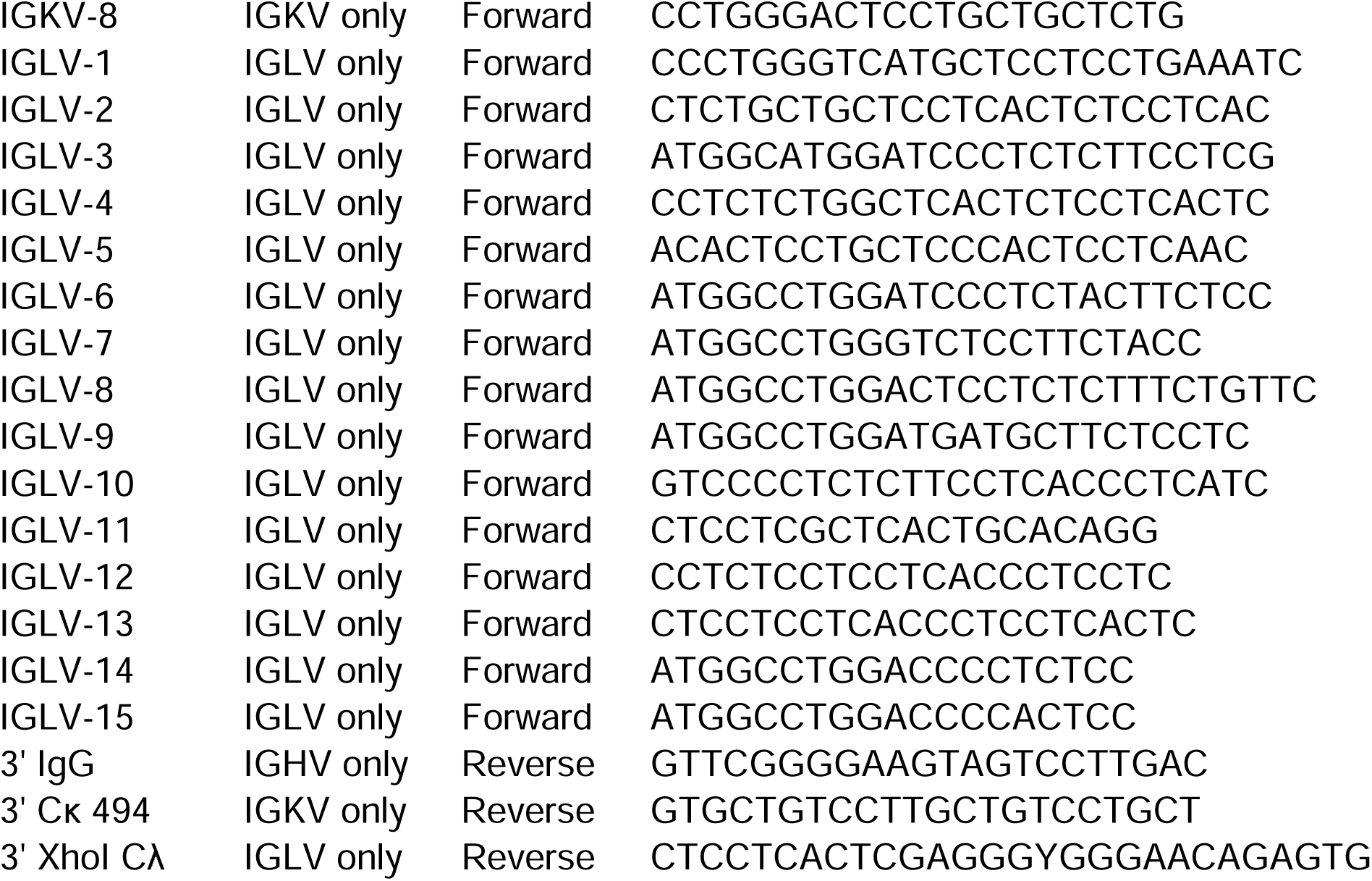
Primers for nested multiplex PCR of IGHV, IGKV, and IGLV loci Primer sets for each of the two reactions in the nested multiplex PCR.

**Supplemental Table 2:**
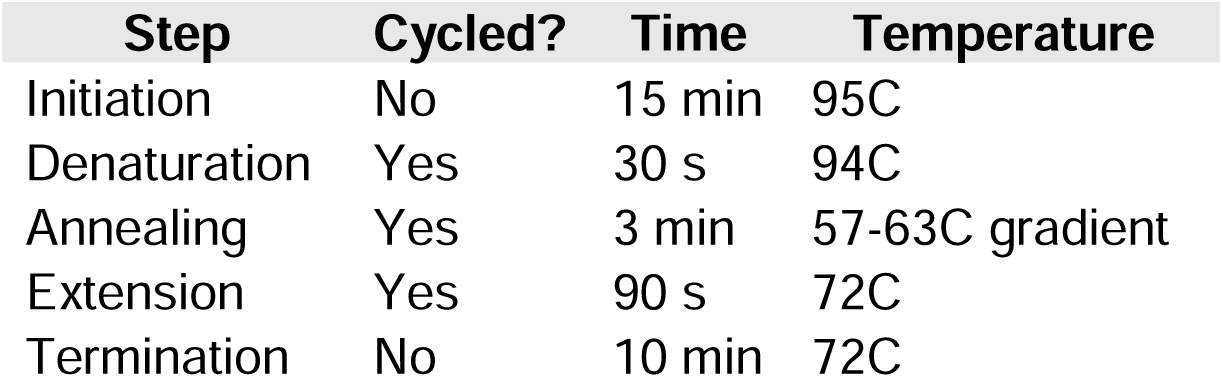
Thermocycling protocol for PCR PCR. Thermocycling protocol. The reaction was run for 30 cycles.

The PCR assay protocol was optimized by testing different concentrations of template DNA in the first amplification attempt with the wastewater DNA extract from Site A and each control. Reactions were run with 1 ug, 300 ng, 200 ng, 100 ng, 50 ng, or 25 ng of template DNA. PCR product from the first nested reaction was cleaned up with the QIAQuick PCR Purification Kit (Qiagen), and its purity and concentration was determined by Nanodrop (Thermo Scientific). Cleaned up PCR product (25 ng) from the first nested PCR was then used as template for the second reaction. The clean-up step was repeated after the second reaction, and the purity and concentration were re-measured. Amplification of DNA in the expected size range of the gene targets (∼500 bp for IGHV and ∼450 bp for IGLV or IGKV) was assessed by gel electrophoresis. Briefly, 16 uL PCR product was pipetted with loading dye into 1% (w/v) agarose gel, and run at 120V in Tris running buffer for 1 hour. Gels were imaged under UV light and the presence of bands at the expected size was confirmed before the wastewater and PBMC positive control amplicons were sent for sequencing (Genewiz).

### 4. Amplicon sequencing

Sample QC, DNA amplicon library preparations, and sequencing reactions were conducted at GENEWIZ, LLC./Azenta US, Inc (South Plainfield, NJ, USA). First, amplicons were quantified using a Qubit 3.0 Fluorometer (Life Technologies), and DNA integrity was checked using a D5000 Screen Tape (Agilent Technologies). NEBNext® Ultra™ II DNA Library Prep Kit for Illumina (New England Biolabs), clustering, and sequencing reagents were used throughout the process following the manufacturer’s recommendations. Amplicons were end-repaired and adenylated. Adapters were ligated after adenylation of the 3’ ends. The DNA library was validated using TapeStation (Agilent Technologies) and was quantified by qPCR. They were then clustered in a flow cell and loaded on the Illumina MiSeq instrument according to the manufacturer’s instructions. The samples were sequenced using a 2×300 paired-end (PE) configuration. The MiSeq Control Software (MCS) conducted image analysis and base calling. Raw sequence data (.bcl files) generated by the sequencer were converted into fastq files.

### 5. Sequence alignment to reference databases

We first compiled three distinct databases of antibody sequences to serve as reference sequences against which we could compare our own amplicon sequences for similarity. To do this, we downloaded all reference sequences for the IGHV, IGKV, and IGLV alleles present in the International Immunogenetics Information System (IMGT) database (https://www.imgt.org/). We also downloaded two databases of antibody protein sequences, The Patient and Literature Antibody Database (PLABDab) and The Coronavirus Antibody Database (CoV-AbDab), which were obtained from the University of Oxford Protein Informatics Group (https://opig.stats.ox.ac.uk/). Heavy and light chain sequences were extracted and ambiguous amino acid residues were removed. These sequences were back translated with Geneious Prime v2025.2.1 (GraphPad Software LLC) using the weighted Mammalian codon usage table and default settings.

FASTA files of raw read sequences were obtained from Genewiz and loaded into Geneious Prime v2025.2.1 (GraphPad Software LLC) for all processing and alignment steps. Raw reads were processed with the following steps: (1) read paired-ends were merged with the BBMerge v38.84 algorithm; (2) reads were trimmed with the BBDuk adapter/quality trimming v38.84 algorithm to remove all Truseq, Nextera, and PhiX adapters and low quality reads (Q<20); (3) duplicate reads were removed with the Dedupe v38.84 algorithm; and (4) reads ≥ 400 bp in length were extracted for downstream analysis (to remove partial sequences). All processing steps were run with default settings. Processed reads were then mapped to each of the three databases using the Geneious mapping algorithm with default settings and set to map all reads to the reference(s) with the highest alignment score and a minimum read-reference overlap of 200 bp (≥ 50% of read aligning to reference). Read alignments were exported as FASTA files. Data analysis and visualization of FASTA alignments were performed with RStudio v2025.05.0 Build 496 (Posit Software, PBC) running R v4.5.0 (The R Foundation) with packages tidyverse, stringr, dplyr, readxl, and seqinr.

## Results

### 1. Amplification of target antibody genes from wastewater

Wastewater from three catchment sites was collected in different cities and regions. Biosolids from each of these sites were collected, lysed, and a raw extract of their DNA was purified (see **Methods**). This DNA served as the template for a nested multiplex PCR targeting three distinct antibody gene loci: the immunoglobulin heavy chain variable region (IGHV), the immunoglobulin kappa variable cluster (IGKV), and the immunoglobulin lambda variable cluster (IGLV). **Figure 1** shows how these regions undergo V(D)J recombination to make unique heavy and light chains. The primers for the nested PCR for these regions are shown in **Supplemental Table 1**. Amplification of DNA of the expected length (∼500 bp for IGHV and ∼450 bp for IGLV or IGKV) was confirmed by gel electrophoresis (data not shown) before amplicon sequencing. The complete methodological approach, including downstream computational assessment, is shown in **Figure 2**.

**Figure 1.**
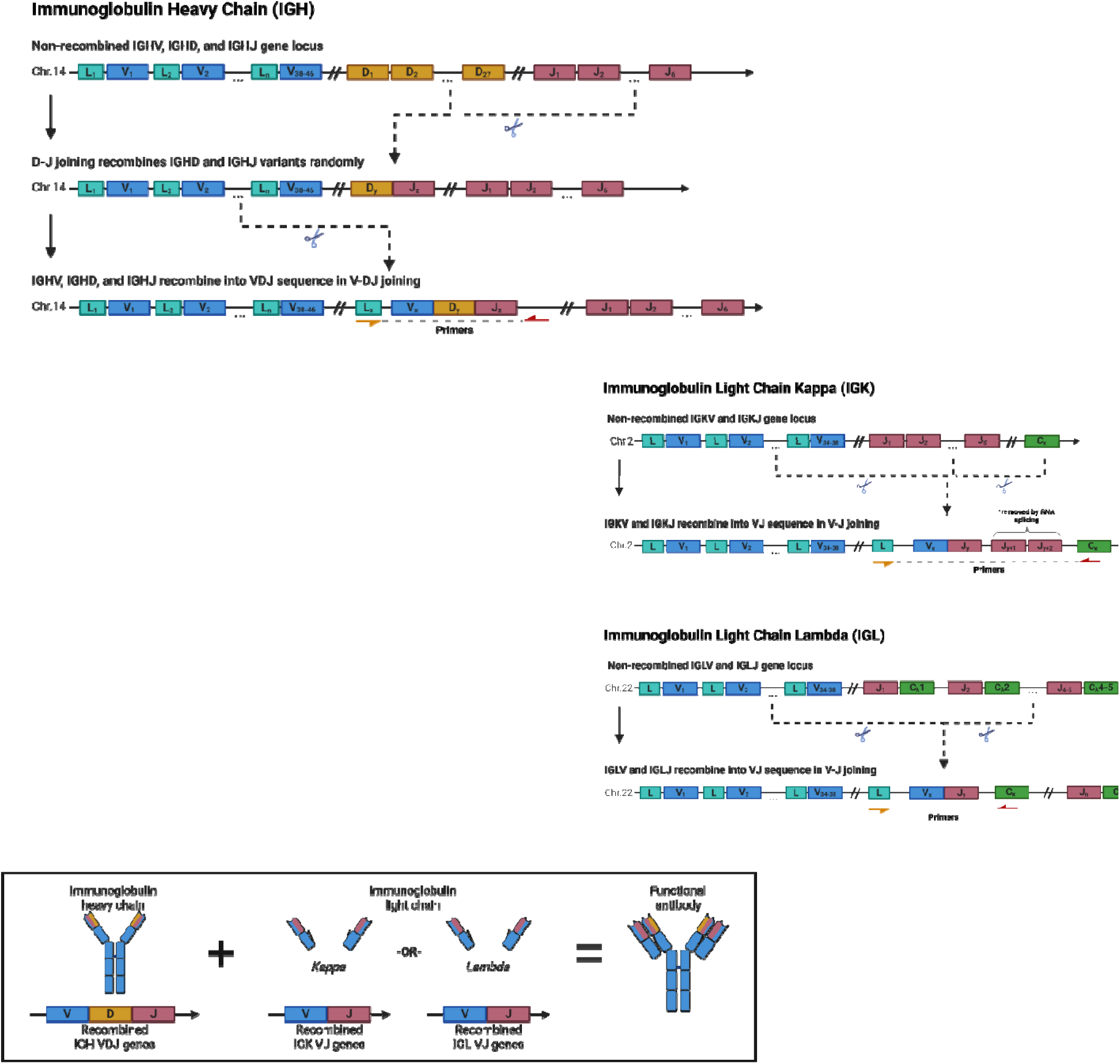
A general overview of V(D)J recombination for each gene locus (IGHV, IGKV, and IGLV) is shown on top. The primer targets are depicted below the final recombinant V(D)J sequences for each locus

**Figure 2.**
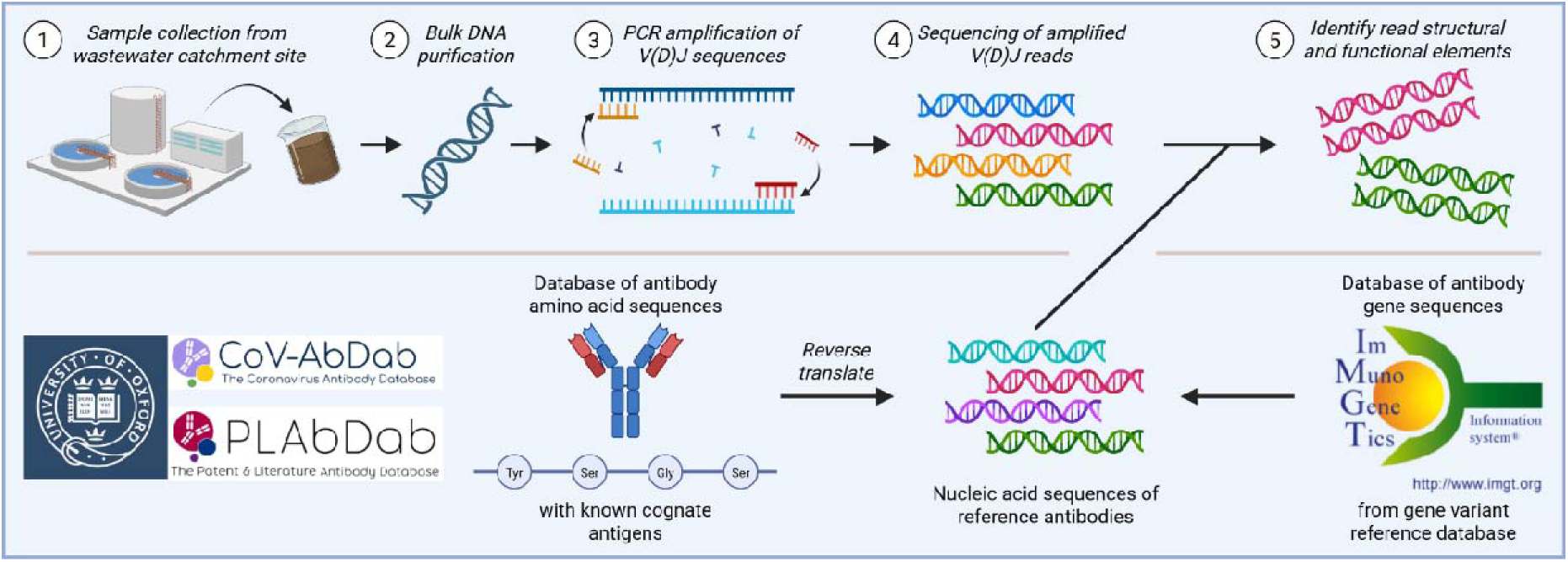
The methodological pipeline for sample processing, PCR amplification/read sequencing, and bioinformatic analysis is depicted below.

Amplicon sequencing produced an average read depth in the hundreds of thousands (**Figure 3**, dark blue). However, we noted that many amplicons were shorter than the expected length, including many that were < 100 bp. “Partial” sequences of < 400 bp were filtered out since full-length reads would be required to span the different structural elements of the gene sequences in question (including the various framework and complementarity-determining regions) and produce the highest-resolution alignments to the reference. Restricting the read pool to only “full-length” reads (≥ 400 bp) reduced the read counts by roughly an order of magnitude (**Figure 3**, blue). These sequences were then aligned to each of the three databases, and the number of reads that aligned to at least one reference from any of the databases were counted (**Figure 3**, light blue).

**Figure 3.**
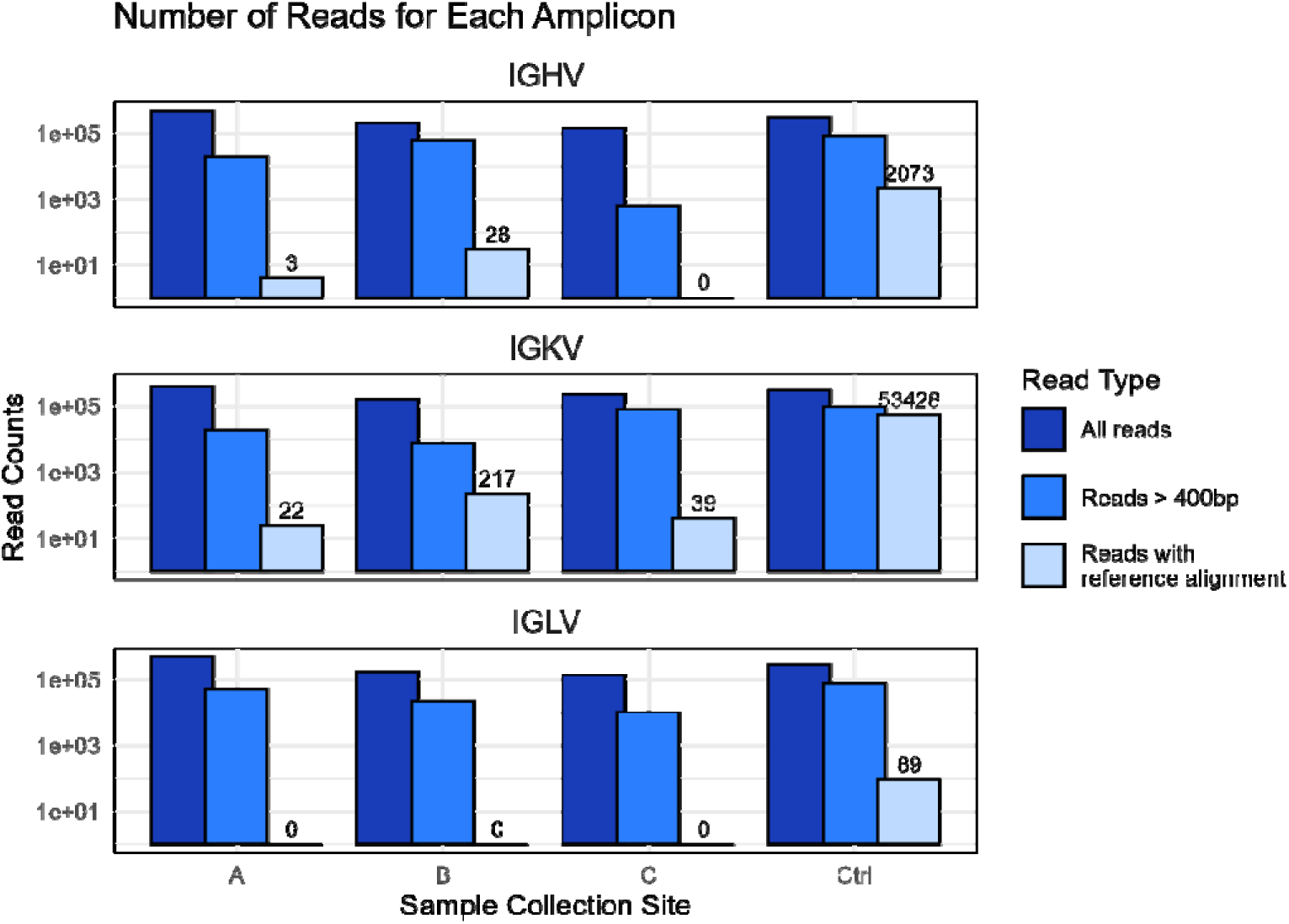
IGHV (top), IGKV (middle), and IGLV (bottom) amplicon read counts are shown for each wastewater site and the PBMC sample control. Reads are counted with the following set of criteria: all sequenced reads (dark blue), reads ≥ 400bp (blue), or reads ≥ 400bp that align to at least one reference sequence with ≥ 50% overlap (light blue).

Numerous reads that aligned to reference sequences from all three databases, including for both heavy and light chain loci. The positive control of amplified DNA from peripheral blood mononuclear cells (PBMCs) produced a greater number of aligned reads at each locus. This highlights the importance of sample/template DNA quality and target sequence abundance for sequencing depth in wastewater samples. The wastewater reads from the light chain PCRs aligned exclusively to the IGKV, and not the IGLV, locus. Although the reason for this is unclear, the higher overall utilization of kappa light chains by B cells may account for some of this discrepancy. For example, it is estimated that kappa light chains constitute roughly two-thirds of the human antibody repertoire^26^, and are up to 20 times more common than lambda chains in mice^26,27^.

In general, some variance in the number of reads aligning to the reference databases across each of the wastewater sites was observed, despite a fairly consistent total read count. Site B produced the greatest number of aligned reads at both heavy (IGHV) and light (IGKV) chain loci, whereas Site C only amplified reads from the IGKV locus. The reasons are not known. It can be speculated that the heterogeneity seen in pathogen nucleic acid signal, likely driven by wastewater composition, applies equally to human nucleic acid.

### 2. Wastewater amplicons elucidate antibody gene variant utilization

After identifying full-length wastewater reads that aligned to some reference sequences, it was next determined which alleles we were detecting at each of these loci (**Figure 4**). To do this, we used the Geneious Prime v2025.2.1 (GraphPad Software LLC) alignment algorithm to map our reads to the International Immunogenetics Information System (IMGT) database of antibody gene reference sequences (IGHV, IGKV, and IGLV genes). The analysis only included alignments with ≥ 200 bp sequence overlap. Furthermore, each read aligned only to a single reference with the best match. For reads that aligned equally well to more than one reference sequence, all alignments were included (thus, the number of alignments is sometimes higher than the total number of reads for a given sample). Identifying the source alleles for the V(D)J recombinant amplicons hints at the clonal diversity of the samples, since a diverse repertoire of antibody sequences would have sequences with many different alleles. Indeed, a high degree of allele diversity in the PBMC control amplicons was observed, particularly at the IGHV and IGLV loci. The IGKV locus, which has the greatest allele diversity for wastewater reads, shows the least allele diversity in the PBMC control sample. This may simply be explained by the greater number of total wastewater reads at this locus compared to IGHV, making the sample look comparatively diverse. Nonetheless, the different IGHV and IGKV alleles detected suggest that the reads represent distinct antibody sequences from different clonal lineages rather than a population of clonally expanded sequences from a singular source.

**Figure 4.**
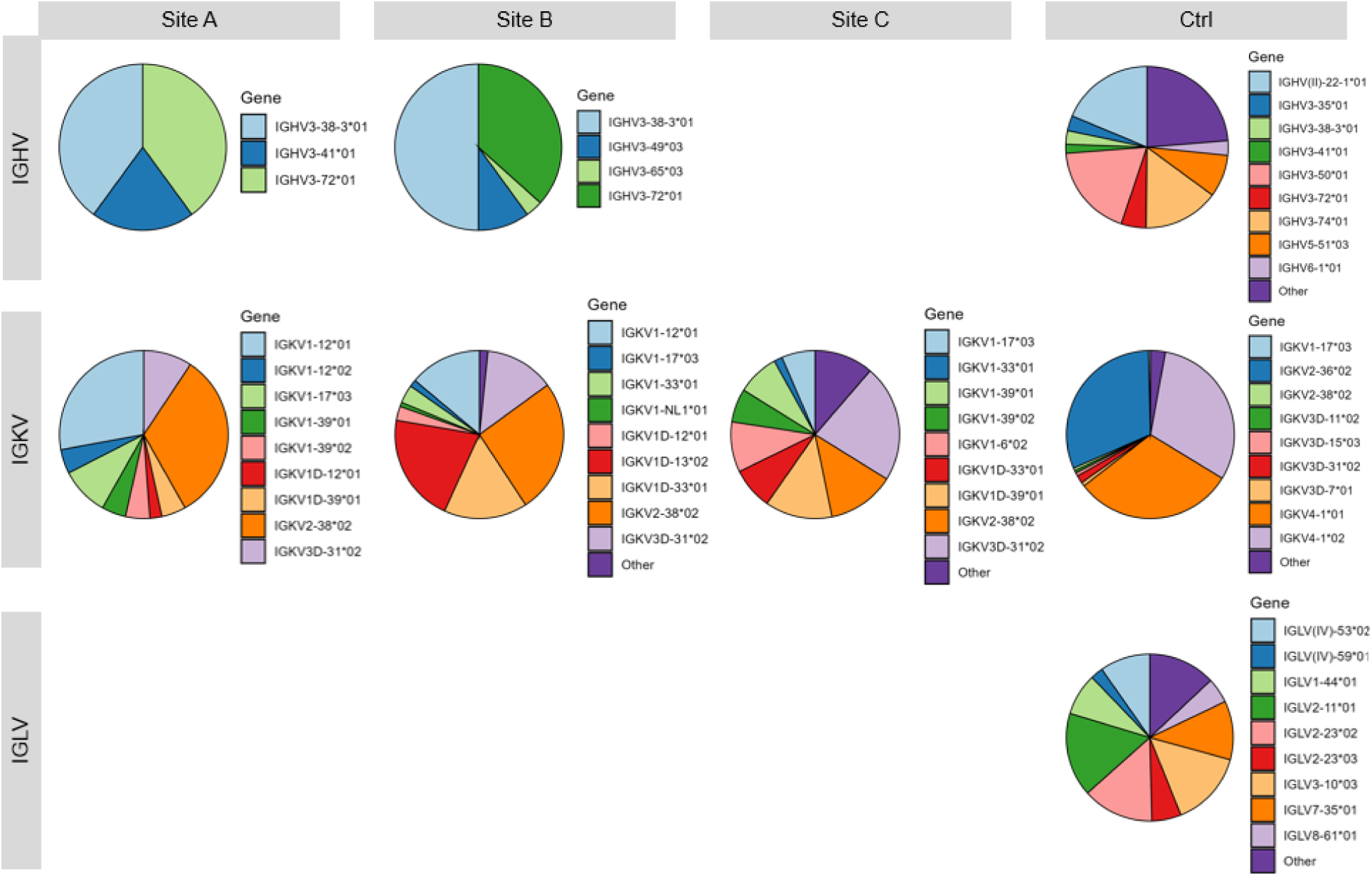
The representation of different alleles from read alignments to the IMGT database are shown for each locus (y-axis) and by wastewater collection site or PBMC sample control (x-axis). Samples with no reads aligning to reference sequences for a given locus are not represented (blank).

Many alleles were identified at relatively high abundance across more than one sample, such as the IGHV3-38-3*01 and IGHV3-72*01 discussed above. We found this notable because the database of IMGT references included 502 reference alleles for the IGHV locus, 121 reference alleles for the IGKV locus, and 155 reference alleles for the IGLV locus. It is reasoned that this could be either because some alleles are utilized preferentially during V(D)J recombination (a biologic phenomenon)^28,29^, or because our PCR primer list is biased towards specific allele sequences (an experimental phenomenon), or some combination of the two.

### 3. Wastewater reads align to heavy and light chain reference sequences

After evaluating which reference alleles the wastewater reads aligned to best at each locus, it was next evaluated how closely related the read sequences were to their best references in the IMGT database of alleles. **Figure 5** shows the alignment for each IGHV heavy chain read below the IGHV reference to which that read was best aligned, where white space along the alignment bar indicates the nucleotides are shared between read and reference at a given position, and a black line indicates there is a gap or mismatch that does not align between the read and reference. The positions of the read start and end points along the reference sequence are denoted below each alignment bar.

**Figure 5.**
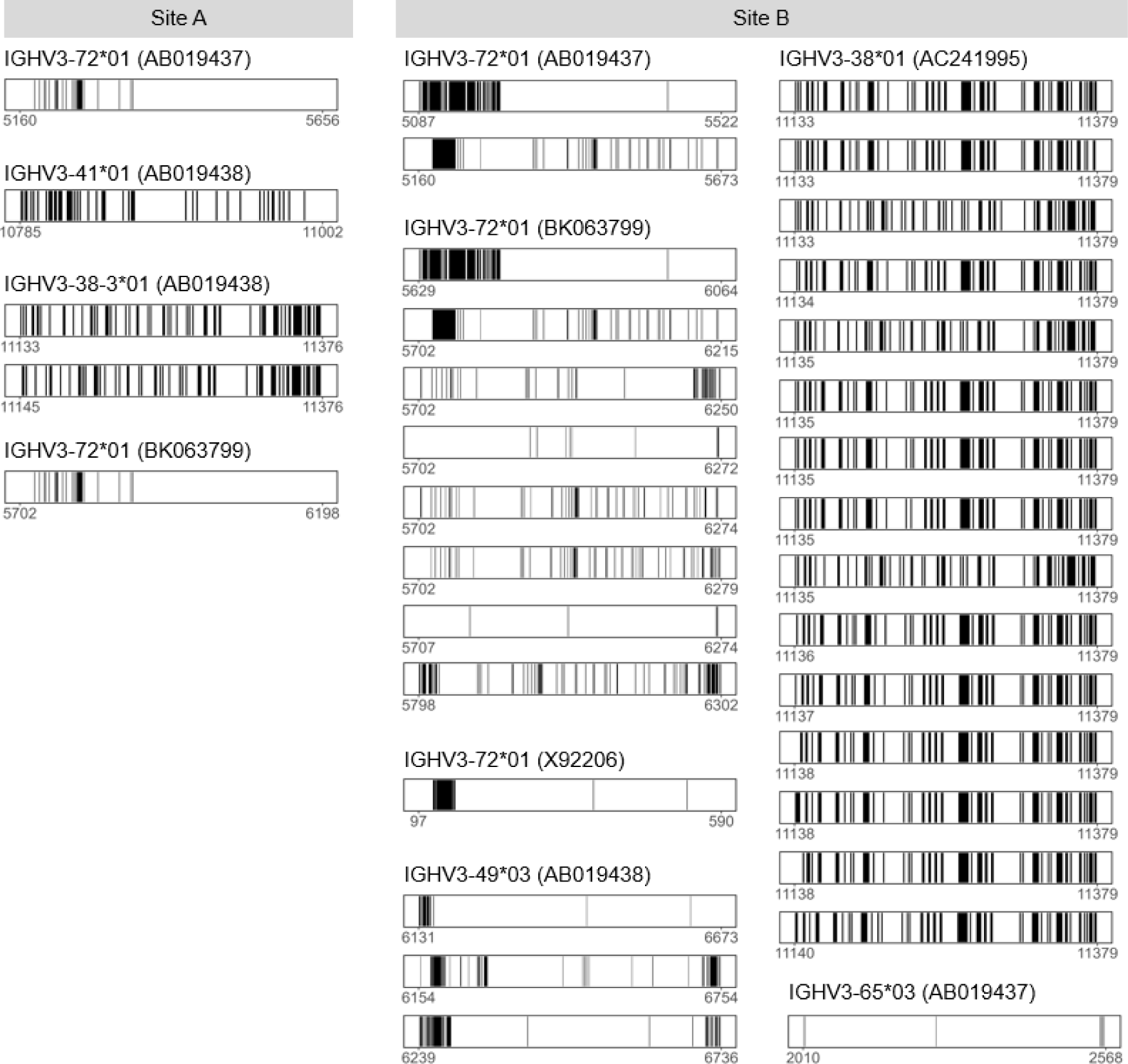
Alignments of each IGHV read to its corresponding reference are shown by wastewater collection site. Nucleotides that match the reference sequence at a given position are shown in white. Read mismatches or gaps are shown in black. The position of the nucleotide along the reference sequence is shown below each read alignment’s x-axis. Read sequence overhangs that map off the 5’ or 3’ end of the reference sequence are not shown here.

Given the significant length of many allele reference sequences, it was also validated that the reads aligned to coding regions of the sequence, which are annotated with structural elements, as most of the references’ sequence space corresponded to 5’ and 3’ UTRs. Indeed, the reads aligned with the 5’ end at the Leader sequence upstream of the V(D)J recombinant and spanned both framework and complementarity-determining regions (CDRs) of the references. However, not all sequences reached the end of full-length reference to the CDR3 region, which exerts the greatest influence on antigen-binding functionality on the antibody of all regions. This is because the filtering parameters only required ≥ 200 bp of the read to align to the reference for the alignment to be extracted, and in some instances nearly half of the read mapped off the 3’ or 5’ end of the reference. In these cases, the read alignments are generally < 300 bp.

However, a number of full-length reads that covered all three CDRs of the reference sequence were identified, including some alignments which are remarkably similar to their reference sequence. For example, the alignment of the Site B IGHV read to IGHV3-65*03 yielded 98.7% identity, with only 6 mismatches over 558 bp. At the lower end, the percent identity for the Site B IGHV reads aligning to the IGHV3-38*01 reference averaged at only 70.7%, suggesting much greater sequence divergence. This may be reflective of the diversity in the human population since it is expected thousands of individuals contribute to the sample.

Interestingly, many of the reads that align to the same reference sequence have a completely different set of mismatches compared to other reads aligning to the same reference sequence. This would suggest that even within a given allele assignment, there is inter-individual sequence variation diversity amongst the read sequences. Meanwhile, other sequences (such as the Site B IGHV3-38-3*01 reads) are distinguished by only a small number of mismatches between each other (no two reads have identical sequences) and could be related or amplified from the same strand of DNA.

The analysis was repeated to produce alignment maps for reads aligning to the IGKV light chain locus references (**Figure 6**). Due to the order of magnitude increase in the number of aligned sequences obtained at this locus, here only one representative read alignment for each unique reference allele is shown. Many of the alleles had numerous reads aligned with varying degrees of similarity, similar to the IGHV alignments in **Figure 5**.

**Figure 6.**
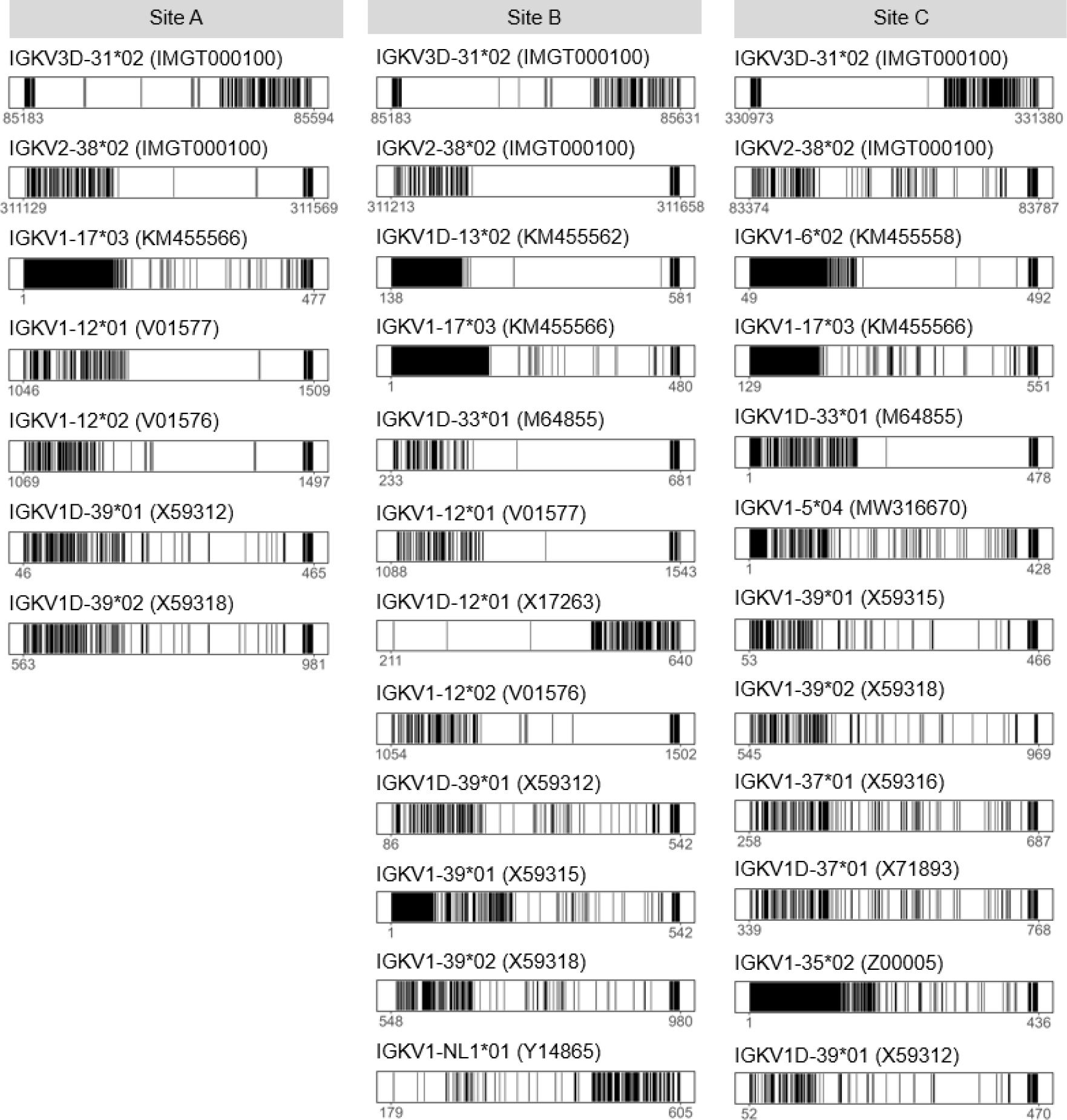
A representative alignment of an IGKV read is shown for each unique reference allele. Reads are sorted by their respective wastewater catchment site. Nucleotides shared between read and reference at a given position are shown in white. Read mismatches or gaps are shown in black. The position of the nucleotide along the reference is shown below each alignment’s x-axis. Read sequence overhangs that map off the 5’ or 3’ end of the reference are not shown.

In both **Figures 5** and **6**, it is noted the higher concentration of gaps and/or mismatches at the 5’ end of the sequence (e.g. for IGHV3-72*01), which is annotated with an intron sequence situated between the Leader sequence and the first framework region encoding the N terminus of the heavy chain protein. A number of the reads also show a higher concentration of gaps and/or mismatches at the 3’ end of the sequence, where the CDR3 region joins the D and J regions of the VDJ sequence. This would be expected, since the process of V(D)J recombination produces the greatest diversity at this junction due to its role in antigen-binding. By contrast, the framework regions in the middle of the sequence show the greatest conservation, which we expected due to their function of providing the antibody’s structure.

### 4. Wastewater reads align to antibody references with known cognate antigens

In addition to looking at wastewater read similarity to the IMGT references, it was also explored whether the wastewater reads could align to a database of antibodies with known functions (i.e. defined cognate antigens). To do this, the University of Oxford Protein Informatics Group’s Patient and Literature Antibody Database (PLABDab) and The Coronavirus Antibody Database (CoV-AbDab) was downloaded. Heavy and light chain amino acid sequences for each antibody were extracted from both databases and reverse translated using the Geneious Prime v2025.2.1 (GraphPad Software LLC) reverse translate algorithm with weighted Mammalian codon usage and default settings. Nucleotide sequence alignments were run as previously described. Reads from both Sites A and B aligned to various references in the PLABDab database. The number of reads aligning to each reference, the percentage of each reference covered by reads, the average percent identity of the aligned reads, and the references’ cognate antigen(s) are shown in **Table 1**. Among the antibodies that can be identified from this first foray, strong evidence was noted for antibodies to human, pathogen (lung bacterium P. aeruginosa) and food (peanut) antigens. Importantly, only the IGHV reads aligned to the PLABDab (heavy chain) sequences, and neither IGKV nor IGLV amplicons aligned to any of the PLABDab light chain sequences in the database. This may be in part because the recombined IGKV-J sequence amplified by PCR includes additional IGKJ sequences that are removed during RNA splicing (**Figure 1**), and thus are present in amplicon sequences but are not reverse translated in the antibody reference sequences.

**Table 1:**
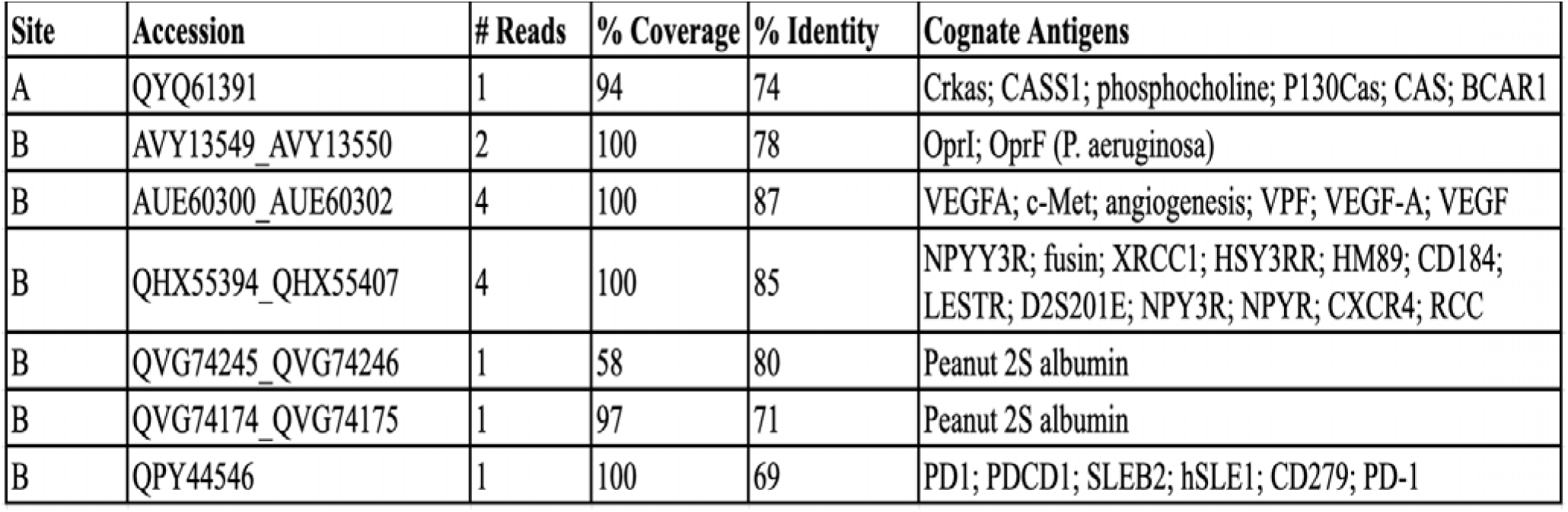
PLABDab reverse translated heavy chain antibody sequences with wastewater IGHV read alignments.

Next, the data was processed to generate representative alignment bars for each of the references to which any wastewater read aligned (**Figure 7**). Because the alignments were generated from reverse translated sequences, they were also compared the amino acid sequences between the original heavy chain sequence from the PLABDab reference to the translated wastewater read sequence. To do this, each read was trimmed to include only the aligned region of the read, excluding the Leader and intron non-coding sequences upstream of the heavy chain protein in these amplicons. These trimmed read sequences were then translated into an amino acid sequence and aligned to the reference. The amino acid alignment for a given read-reference pair is shown below each nucleotide alignment. Only alignments that spanned the entirety of the reference sequence are shown here. The process was repeated for the CoV-AbDab reference database, shown in **Figure 8**.

**Figure 7.**
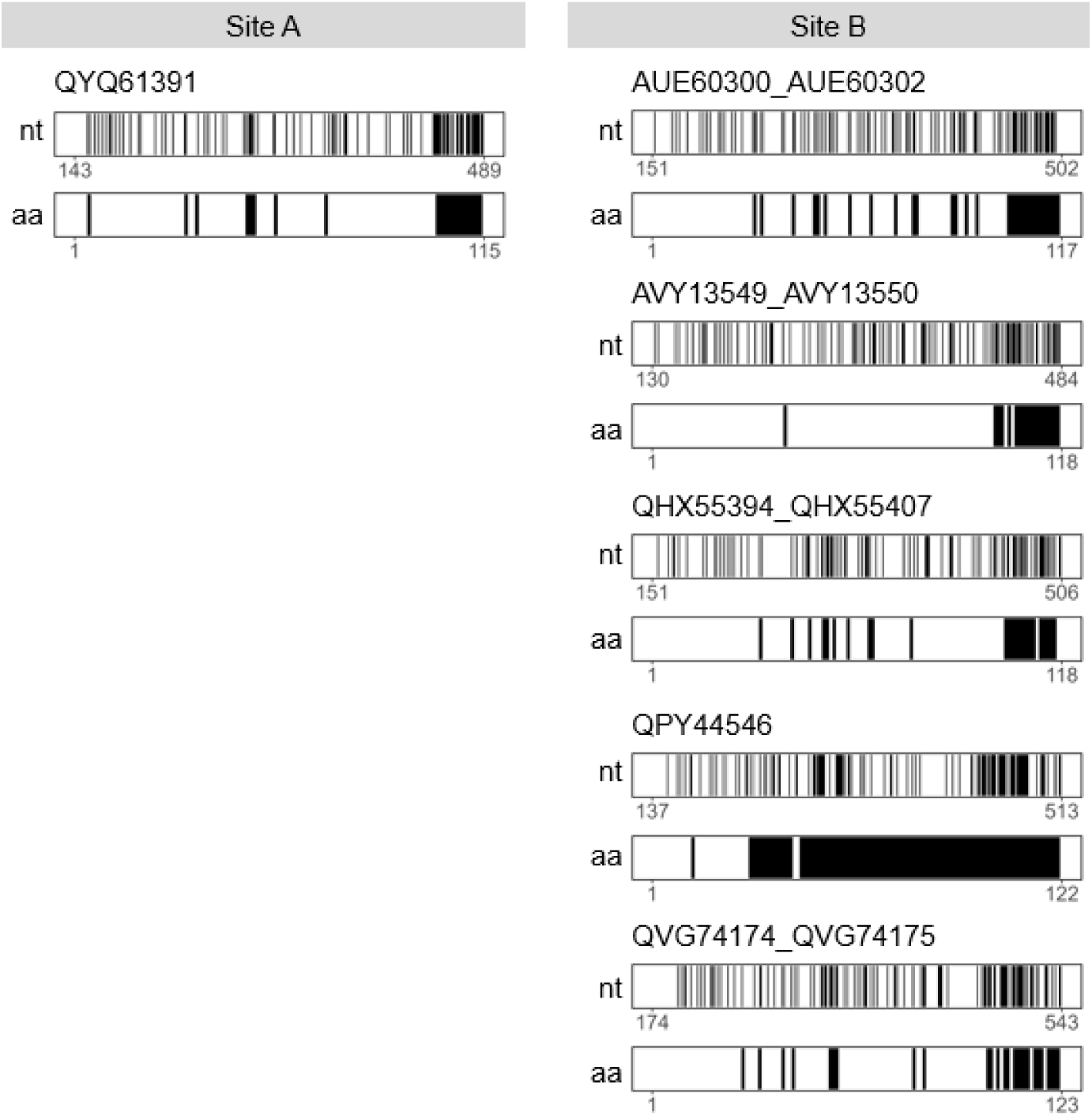
A representative alignment of a wastewater read is shown for each PLABDab antibody heavy chain reference sequence to which a read was aligned. Each alignment is shown for both nucleotide (nt, top) and amino acid (aa, bottom) sequences. Reads are sorted by the catchment site from which they were amplified. Nucleotides or amino acids that match their reference sequence at a given position are shown in white. Mismatches or gaps are shown in black. The position of the nucleotide or amino acid along the read sequence is shown below each alignment’s x-axis. Read sequence overhangs that map off the 5’ or 3’ end of the reference sequence are not

**Figure 8.**
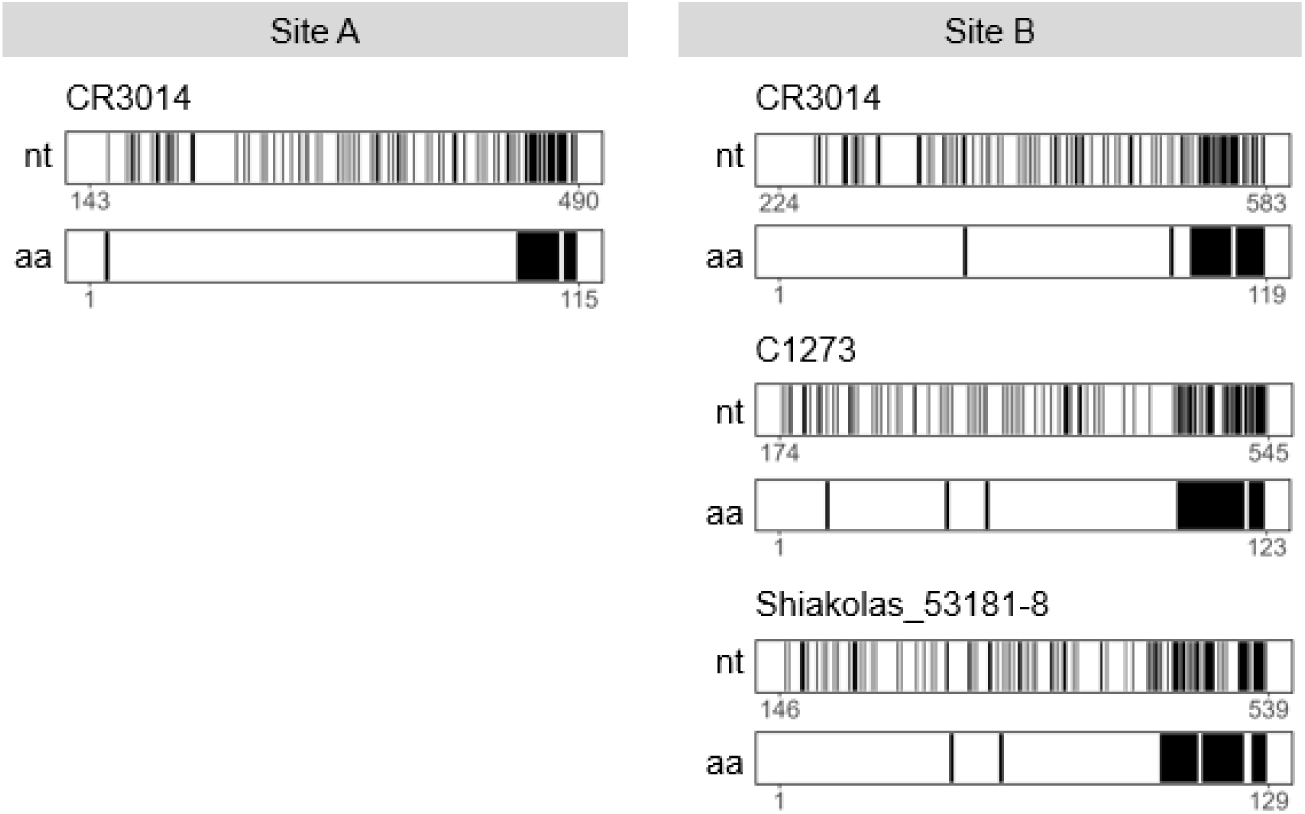
A representative alignment of a wastewater read is shown for each CoV-AbDab antibody heavy chain reference sequence to which a read was aligned. Each alignment is shown for both nucleotide (nt, top) and amino acid (aa, bottom) sequences. Reads are sorted by the catchment site from which they were amplified. Nucleotides or amino acids that match their reference sequence at a given position are shown in white. Mismatches or gaps are shown in black. The position of the nucleotide or amino acid along the read sequence is shown below each alignment’s x-axis. Read sequence overhangs that map off the 5’ or 3’ end of the reference sequence are not shown here.

Many of the nucleotide alignments contain many more mismatches than the amino acid sequence alignments. We suspect that this may be caused by the *in silico* reverse translation data processing, which probablistically assigns codons based on weighted Mammalian codon usage tables. Thus, the nucleotide sequence of the reference may not reflect the actual codon usage observed in IGHV, IGKV, or IGLV loci. By contrast, the amino acid sequence alignments reflect the actual antibody protein produced by both sequences, and are therefore more relevant.

However, it is important to include the nucleotide alignments because of the presence of deletions, insertions, or gaps in the sequences, which can cause a frameshift and make the alignment appear artificially discordant. The amino acid alignment for the QPY44546 reference sequence, for example, shows poor sequence agreement despite an average number of mismatches in the nucleotide alignment. This is because there is a single nucleotide deletion in the read sequence relative to the reference, which introduces a frameshift that changes the rest of the amino acid sequence downstream.

We also noted that the amino acid mismatches are concentrated at the end of the sequence, particularly the final 8-12 amino acids. This portion of the sequence corresponds to the CDR3 region of the heavy chain which determines the antibody’s antigen-binding function and suggests that although the wastewater reads differ from the reference to which the reads are aligned, the read sequence does encode the conserved framework region that supports the structure of the heavy chain. The importance of this finding is that with improvement of the methods and analysis associated herein, the amino acid regions of this chain can be readily determined at much greater resolution.

## Discussion

To our knowledge, this is the first report of human antibody genes sequenced from wastewater, and, importantly, the first reported nuclear genome analysis from wastewater. Targeted PCR and sequencing resolved allelic usage across these genes and matched the resulting sequences to databases of antibodies with known specificities. Although preliminary, the results provide proof-of-principle that wastewater may be used to monitor population immunity to pathogens through the amplification of antibody repertoire regions.

Human genetic material has been recovered from wastewater before but exclusively at mitochondrial, not nuclear, loci. Kapoor and colleagues amplified and deep-sequenced the human mitochondrial hypervariable region II from sewage-impacted surface water as a fecal source-tracking marker^30^, and later work recovered human mitochondrial DNA from shotgun metagenomes of municipal sewage across forty-four cities to estimate population haplogroup composition^31^. More recently, human genomic material has been shown to be an unavoidable component of environmental DNA sampling generally, sufficient in quality to support variant calling^32^. In parallel, a separate literature has established that human immunoglobulins (the proteins themselves) are present and functional in wastewater: pathogen-specific anti-SARS-CoV-2 IgG and IgA were first demonstrated by ELISA^22^, and a subsequent study characterized the wastewater antibody pool as predominantly secretory IgA of mucosal origin, retaining affinity for SARS-CoV-2 and influenza A antigens and varying with the respiratory season^23^. That study identified immunoglobulin variable-region peptides by mass spectrometry, establishing that variable domains survive in wastewater. However, peptide mapping against a reference proteome cannot resolve the features that define a repertoire (the V(D)J recombination junction, CDR3 identity, clonal lineage, or somatic hypermutation (because tryptic peptides assigned to a variable region are matches to germline consensus rather than to clonotypes). No study to date has sequenced the rearranged immunoglobulin genes themselves from an environmental sample, and no human nuclear protein-coding locus has been targeted and sequenced from wastewater. Here we address both gaps.

Despite our success in amplifying and characterizing antibody gene sequences from wastewater, we also acknowledge the fundamental limitations of this data for informing public health decision making. First, antibody sequence data cannot definitively determine the in vivo function (if any) of the encoded antibody or the antigen/pathogen against which the sequence was clonally selected. Although sequence similarity with published antibody genes of known function could theoretically suggest a function, we did not identify any read sequence with striking similarity to a previously published antibody at the CDR3 site which is most important for cognate antigen binding. As antibody sequence databases with functional annotations continue to expand, this limitation may be diminished over time. However, given the estimated 10^12^ - 10^15^ theoretically possible antibody sequences^33,34^, it is unlikely that such a database would ever be comprehensive enough to match entire repertoires to cognate antigens. To this end, a number of machine learning algorithms are being developed to predict cognate antigen binding based on the nucleotide sequence^35–38^. Further research is needed to validate and improve the accuracy of these models.

A second limitation of our approach is the unpaired bulk sequencing of heavy and light chains from wastewater. Because both chains are involved in antigen binding function, even if a heavy and light chain from the sample wastewater composite both matched with a paired antibody sequence in our databases, it could not be stated with certainty that they were derived from the same B cell clone within that sample. Additional computational tools could be used to assess the likelihood of heavy and light chain pairing for two sequences^39,40^, but only single-cell based sequencing approaches could definitively assign heavy and light chain pairs. This approach would limit the throughput and sequencing depth in the wastewater testing pipeline but would have the added benefit of enriching target cell populations (e.g. human B cells) from the wastewater to increase the relative abundance of the target DNA and improve the overall quality of the sequencing data.

Thus, to translate wastewater antibody repertoires into actionable public health insights, several things need to happen. Crucially, cognate antibody-antigen databases must continue to expand, empowered by next-generation sequencing technologies. This will enable better alignments with wastewater reads, but it will also facilitate the development of more accurate machine learning models. And finally, greater sequencing depth will need to be achieved in order to comprehensively evaluate the immunological status of a population against a range of antigens and pathogens of interest, and to quantify the relative strength of a given signal. Only then can the nucleotide signal be correlated with transmission of the cognate pathogen in the community. Towards this end, we confidently report the amplification and sequencing of 31 IGHV and 278 IGKV reads from 3 wastewater catchment sites. As such, our study provides a first step towards the monitoring of population immune status by establishing a high-throughput and unbiased system for obtaining amplified human antibody gene sequences from wastewater. We hope that future research will enable a new generation of wastewater-based epidemiology, wherein public health officials can proactively assess a community’s vulnerability and preparedness to various pathogens and propose tailored interventions to prevent outbreaks of infectious disease.

We would be remiss not to state the obvious implication of this work. If rearranged immunoglobulin genes can be amplified and sequenced from municipal sewage, then so, in principle, can any human gene. This matters because the ethical protections that currently justify wastewater-based epidemiology rest almost entirely on the premise that pooled community samples are inherently anonymous. That premise holds well for viral genomes and for mitochondrial haplogroups, which describe populations rather than persons. We therefore offer this report not only as a technical advance but as a call for the field to establish governance before capability outpaces it *i.e.*, limits on what human loci may be sequenced from community samples, requirements for catchment-level aggregation and site anonymization, prohibitions on linkage to any other dataset, and community engagement in sewersheds where surveillance is conducted. Wastewater surveillance has earned public trust because it watches pathogens and not people. That trust is an asset worth protecting deliberately while also recognizing the remarkable power for improving population health that this technology and analysis may eventually bring.

## Data Availability

All data produced in the present study are available upon reasonable request to the authors.

## Acknowledgements

This study was reviewed by the Institutional Review Board of Baylor College of Medicine and determined to be exempt from human subjects research oversight as it does not constitute human subjects research since wastewater samples are collected at the level of municipal catchments and contain no identifiable private information about any individual (H-57756). Sampling site identities are withheld throughout this report to further limit the possibility of community-level attribution. The study design, analytic approach, and this manuscript were additionally reviewed and discussed by the Ethics Working Group of the Texas Wastewater and Environmental Biomonitoring (TexWEB) program, whose guidance shaped our approach to data reporting and site anonymization. We thank the municipal water utilities and public health partners who collected and provided the wastewater samples, without whom this work would not be possible.

